# Quality of Cervical Cancer Screening at a Family Health Unit in Salvador-BA

**DOI:** 10.1101/2021.05.31.21257712

**Authors:** Julita Maria Freitas Coelho, Jamilly Souza Da Silva, Letícia Fonsêca Santos, Lyvia Mirelle Carneiro França, Gláucia Alencar Ponte, Lorena Moura De Assis Sampaio, Caroline Ramalho Galvão, Lorena Ramalho Galvão, Bruna Matos Santos Dantas, Sarah Dos Santos Conceição, Érica Velasco Dias Gomes, Maria Emília Cirqueira Silva, Ana Clara Silva Oliveira, Caroline Santos Silva

## Abstract

**Introduction:** Cytopathological examination is the main method for the screening of cervical cancer, its occurrence can be prevented or minimized through screening, with good coverage, quality of collection and analysis and women’s adherence to the examination.

**Objective:** To describe the quality of cytopathological exams performed at a family health unit in Salvador - Bahia during the years 2015 and 2016.

**Method:** A cross-sectional study, of an exploratory descriptive character, was conducted, using a database from a previous study carried out in a health unit in Salvador / BA and approved by the Ethical Committee from State University of Feira de Santana.

**Results:** Preventive gynecological exams of 1,350 women were analyzed, but 330 (24.4%) of them could not be evaluated due to the absence of the report in the health unit and the presence of acellular or hypocellular material that prevented the analysis. From 1020 exams, 45 (3.3%) presented an unsatisfactory sample, without conditions for analysis, while 589 (43.19%) showed only squamous cells. From 392 (29.0%) reports with an adequate study sample, 150 (%) were women with a mean age of 38.49 years. These were located and composed the final sample of the present investigation.

**Final Considerations:** We suggest the need for investments in continuing education for professionals involved in performing cytopathological exams, in order to ensure the right of women to comprehensive and quality health care.

## INTRODUCTION

Cytopathological examination (CE) is the main method for early identification of cervical cancer (CC) and its precursor lesions. Also known as Papanicolau, according to Caetano, Viana e Girianeli (2006), it is recognized as a fast, low cost, and effective for detection, although its vulnerability for collection bias, plate preparation and its interpretation.

Regarding sampling for this assay, its elegibility evaluation has been considered a quality indicator, and the sample that has a representative amount of well distributed, fixed and coloured cells, allowing a diagnostic conclusion, is considered adequate for analysis. The samples may contain squamous, glandular and metaplastic cells (INCA, 2016). On the other hand, samples with an unsatisfactory reading due to technical aspects or cellular sampling leads to an unfeasible diagnosis (BRASIL, 2016).

Most of cervical cancer cases could be prevented by the adherence to cytopathological examination, with a good coverage screening, quality sample collection and correct analysis (GONTIJO et al, 2004). According to the World Health Organization (WHO), with at least 80% of the population screened, with confirmed diagnosis and adequate therapy, there could be a reduction from 60 to 90% of cervical cancer cases. Some developed countries show a reduction of CC incidence of 80% in locations where cytopathological examination was performed with quality and the coverage was reached, and diagnosis was confirmed and adequate therapy was delivered (BRAZIL, 2011).

In order to reduce CC morbidity and mortality in Brazil, the Ministry of Health (MoH) recommends annual examination for sexually active women who are 25 to 64 years old; women younger than 25 years old should avoid this examination. The examination can be performed every three years for women presenting two consecutive normal results, with an interval of one year between them. The first two examinations must be performed annualy, and, if both of results are negative, the next must be taken every three years (BRASIL, 2011; BRASIL, 2016).

Some factors influence on the detection of lesions, such as unsufficient material collection; inadequate smears and plate preparation; technical deficit in cytopathology laboratories; women’s low adherence on being examined (LAPIN et al, 2000). Therefore, the sample analysis can be compromised due to an unsatisfactory collection, invalidating the CE as a screening measure for CC (AMARAL et al., 2008). A false-negative result compromises the patient’s prognosis since the non visualization of suggestive lesions of CC may increase its morbimortality (LEITÃO et al, 2008)

Therefore, the conduction of research regarding the quality of CC screening may offer information that evidence the quality of CE performed, as well as the attainment of parameters to guide strategies to personnel qualification regarding sample collection in CE. The aim of the present study is to describe the quality of cytopathological examinations performed at a Family Health Unity (FHU) in Salvador - Bahia, between 2015 and 2016..

## MATERIAL AND METHODS

We performed a cross sectional, descriptive and exploratory study, using a database from a previous study. The present study was approved by an Ethical Research Committee (n.º 2.548.705).

The research context was based on a FHU from Salvador county, state of Bahia. Data from patient hospital records and registry from CE collection were analysed between december 2017 and june 2018, in order to identiy potential participants. Subsequently, women located by community health agents were invited to participate in the present research. Women that accepted signed a consent form, and were interviewed with semi-structured questionnaire.

Therefore, women registered in the FHU, presenting CE results with squamous and glandullar cells, with or without precursor lesion for CC, and with inconclusive results, were selected for the study. According to elegibility criteria, 29% of women were eligible, and 150 were inclued in the final phase of the study.

Descriptive analysis comprised simple and relative frequencies regarding categorical variables, and central tendency measures for continuous variables. Statistical analyses were performed on Statistical Package for the Social Sciences (SPSS) version 17.0 and STATA version 10.0 softwares.

## RESULTS

Between december 2017 and june 2018, 1,350 cytopathological examinations were performed in San Martin 1 FHU. Nonethless, only 392 reports presented adequate samples, whilst 3.3% had unsatisfactory sample and 43.19% showed only squamous cells. We point out that 24.44% of examinations could not be evaluated because its reports were not available in the health unit, even with a second copy being requested, or did not show a detailed registry (FIGURE 1).

**Figure 1-.**
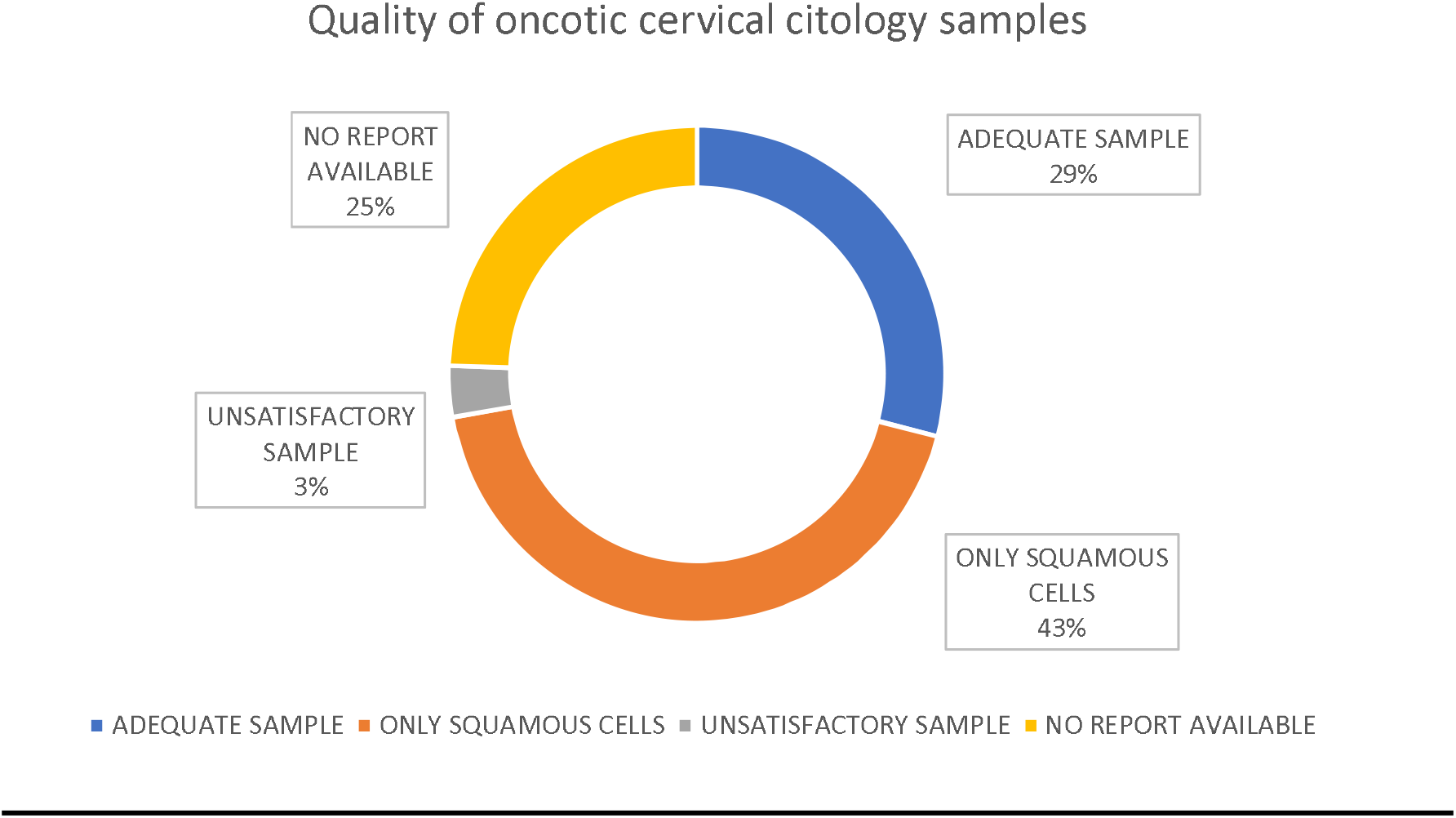
Quality of oncotic cervical citology samples. Salvador, BA, Brazil (n=1,350)

Participants were in average 38.49±13.68 (%) years old (median of 37 years old), and the majority self reported as Black and/or Brown (138/92.0%). Most part of women in the sample did not have a Professional occupation at the moment of the interview (89/59.3%) (TABLE 1).

**Table 1.** Sociodemographic characteristics and lifestyle habits from the studied sample, obtained by the survey form. Salvador, BA, Brazil (n= 150).

| Characteristics | n | % |
| --- | --- | --- |
| **Age** |  |  |
| Mean | 38.49 ± 13.68 |  |
| Median | 37 (18 - 81) |  |
| **Race/skin color** |  |  |
| White/Yellow/Indigenous | 12 | 8.0 |
| Black/Brown | 138 | 92.0 |
| **Education level** |  |  |
| ≥ 8 years | 95 | 63.3 |
| < 8 years | 55 | 36.7 |
| **Occupation** |  |  |
| Employed | 61 | 40.7 |
| Unemployed | 89 | 59.3 |
| **Smoker** |  |  |
| No | 142 | 94.7 |
| Yes | 8 | 5.3 |
| **Alcohol consumption** |  |  |
| No | 71 | 47.3 |
| Yes | 79 | 52.7 |
| **Physical activity** |  |  |
| ≥ 2 days/week | 31 | 20.7 |
| 0 to 1 day/week | 119 | 79.3 |

Regarding lifestyle, there was a prevalence of women consuming alcoholic beverages (79/52.7%), non smokers (142/94.7%) and with low consumption of ilicit drugs (n=5/3.3%). We also observed that the majority of women had a low frequency of physical activity (119/79.3%) (TABLE 1).

Results of reproductive conditions show that the majority of women had the first sexual contact up to 18 years old (102/70,0%), and most of them also referred to relate themselves with up to 4 partners (102/68.0%), and almost all of them had one fixed partner (149/99.3%) (TABLE 2).

**Table 2.**
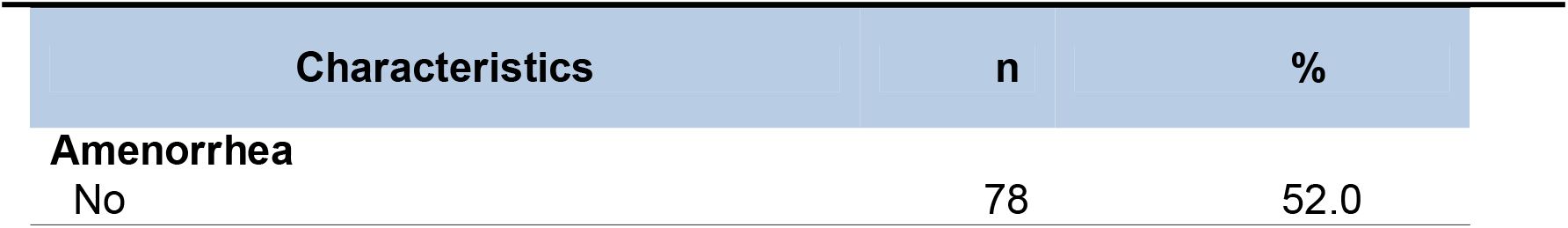
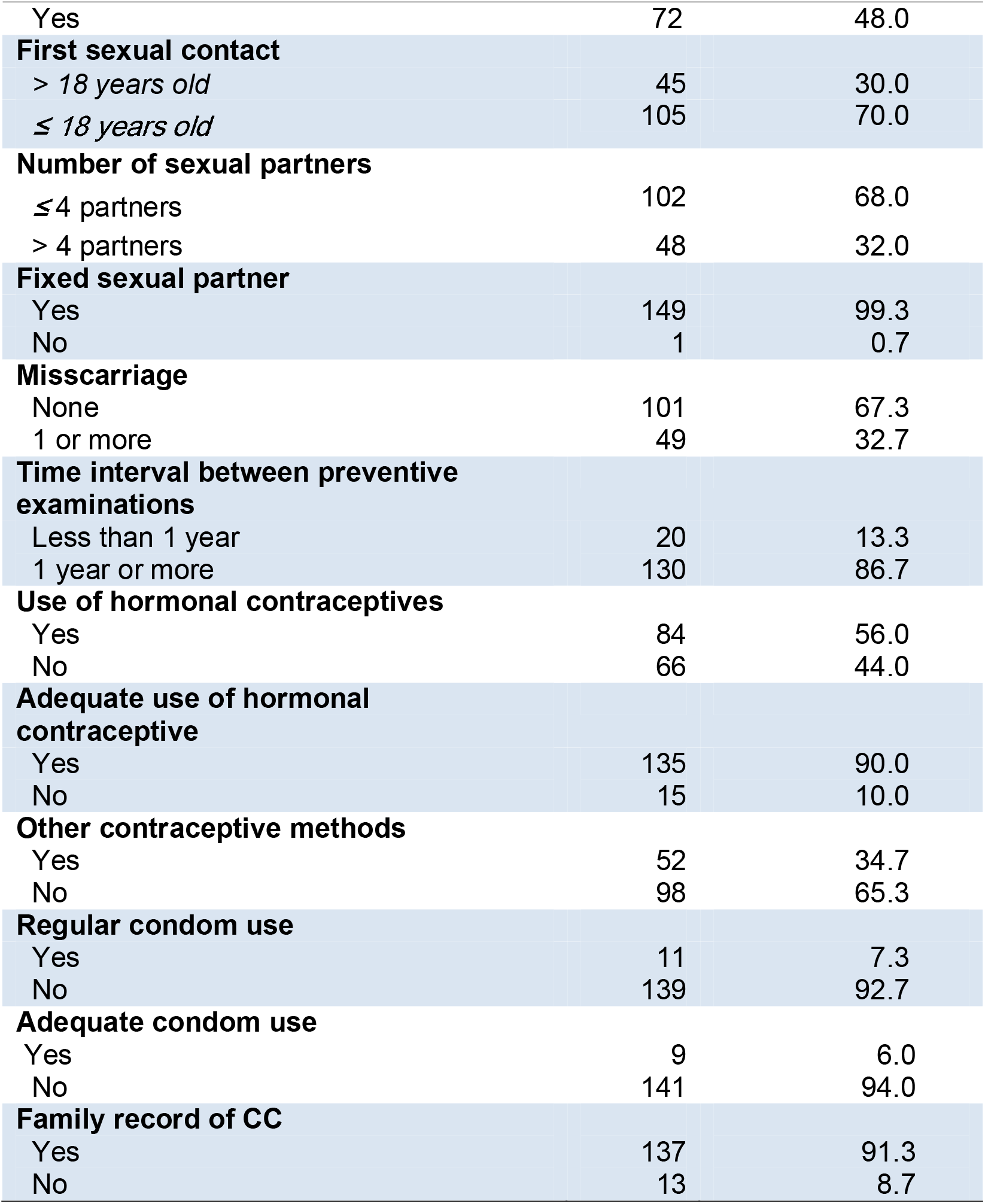
Clinical and reproductive conditions and family records of women included in the studied sample, survey form. Salvador, BA, Brazil (n=150).

The majority of women reported more than one year of interval between the last two gynecological preventive examinations (130/86.7%) and most of them reported using of hormonal contrapectives (86/56.0%) adequately (135/90.0%); similarly, most of women reported they did not use other type of contraceptives (98/65.3%). There were few reports regarding regular (11/7.3%) and adequate use of condoms (141/94.0%) (TABLE 2).

We observed from cytopathological examination records that only a few part of women showed precursor lesions of CC (17/11,2%), where the majority of lesions were Atypical Squamous Cells of Undetermined Significance (ASCUS) (7/4,7%), and the least observed were atypical glandular cells of undetermined significance (1/0,7%). There was also a low frequency of ASCUS in possibly non neoplasic cells, cannot exclude a high grade intraepithelial lesion (HSIL) (2/1,3%), of low grade intraepithelial lesion, including human papillomavirus infections, mild dysplasia and intraepithelial cervical neoplasia I (low grade intraepithelial lesion) (5/3,3%), and high grade intraepithelial lesions (intraepithelial cervical neoplasia II and III, in situ carcinoma) (2/1,3%) (TABLE 3).

**Table 3-.** Aspects of vaginal microbiota and other gynecological characteristics from cytological vaginal examination from women included in the studied sample. Salvador, BA, Brazil (n=150).

| Characteristics | n | % |
| --- | --- | --- |
| **Presence of lesion** |  |  |
| No | 133 | 88.7 |
| Yes | 1 | 11.2 |
| **Cellular changes** |  |  |
| ASCUS | 7 | 4.7 |
| ASC-H | 2 | 1.3 |
| AGC | 1 | 0.7 |
| LSIL | 5 | 3.3 |
| HSIL | 2 | 1.3 |
| Not applicable | 133 | 88.7 |
| **Atrophy with inflammation** |  |  |
| No | 138 | 92.8 |
| Yes | 2 | 8.0 |
| **Vulvar lesion** |  |  |
| No | 144 | 96.0 |
| Yes | 6 | 4.0 |
| **Vaginal lesion** |  |  |
| No | 149 | 99.3 |
| Yes | 1 | 0.7 |
| **Leukorrhea** |  |  |
| No | 67 | 44.7 |
| Yes | 83 | 55.3 |
| **Microbiota** |  |  |
| Physiological | 104 | 69.3 |
| Pathological | 46 | 30.7 |
| **Metaplasia** |  |  |
| No | 148 | 98.7 |
| Yes | 2 | 1.3 |
| **Benign alterations** |  |  |
| Normal | 149 | 99.3 |
| Altered | 1 | 0.7 |
| **Onset of malignancy** |  |  |
| Positive | 23 | 15.1 |
| Negative | 127 | 83.6 |

We also observed the precence of leukorrhea in the most part of examination reports (83/55.3%); pathological microbiota was less frequent than physiological (46/30.7%). Metaplasias were rare (2/1.3%), as well as benign alterations (1/0.7%); however, positivity for malignancy was more frequent (23/15.1%) than the two categories cited above (TABLE 3).

## DISCUSSION

Regarding the initial number of examinations evaluated (n=1,350), only 29% had na adequate collection for diagnosis. Thus, more than 1,000 (71%) women did not have the opportunity to be screened regarding CC or its precursor lesions. That is because the sample was unsuficcient, unsatisfactory or only had squamous cells, therefore, without conditions for an adequate diagnosis.

Moreover, the examination alterations frequency was 11.2%; therefore, there is a possibility that more than 112 women had these alteratioins at the time of sample collection, but were not diagnosed, or did not receive a control measure or treatment for such alterations, and did not have the right to know their real gynecological health status.

On the other hand, the Brazilian MoH recommends a minimmum frequency of unsatisfactory samples of 5% as criteria for quality of collection and sample preservation in the screening examination for CC (BRASIL, 2016). We observed in the present study that the frequency of unsatisfactory samples were according to the Brazilian MoH recommendation, although values close to zero better indicate an adequate screening (BRASIL, 2012).

In contrast, Santos (2011) showed the elevated frequency of unsatisfactory samples on vaginal cytopathological examinations in Brazilian states. The state of Maranhão had the best evidence for showing a frequency of 5.6%, while Rio Grande do Norte had a frequency of 2.0%. Central-western, Southeast and Southern regions had states with a frequency close to the Brazilian MoH recommendation. In contrast, in a study performed at Doresópolis, Minas Gerais, all plates (100%) were considered adequate for cytopathological examination (SILVA; ARAÚJO; ARAÚJO, 2011). Therefore, since sample adequacy is an important indicator of quality in cytopathological examinations (BRASIL, 2016), it is a positive aspect regarding CC screening in the family health unit evaluated in the present study.

This examination is important for the prevention of CC and has a low cost, therefore, the personell qualification is essential and imperative (LEITÃO et al, 2008).

This is probably because the big ammount of unsatisfactory samples may be related to personell accuracy during the cytopathological examination.

Adequate cytopaythological examination is mandatory for ensuring women’s health, which is a major social right protected by the current legal framework (BRASIL, 1988). Social, political and economic policies focused on the reduction of risk for diseases and harms is a State’s responsability. The Unified Health System (Sistema Único de Saúde – SUS) forecasts that access to health must happen in an universal and equitable manner, looking forward to health promotion, protection and recovery (BRASIL, 1990). The above mentioned collection involves mannual work, and, therefore, investments on health professionals qualification is required.

Underpinning with the present study, results from a women’s health research, conducted at Aracaju, Sergipe, showed that only 46.2% (154) of samples were adequate for the analysis. The same study showed that cytopathological examination, cervical oncotic cytolgy test, is considered the best procedure to detect the first lesions, which indicates its systematic undertaking in women who are 25 to 64 years old. Finally, the authors point that social, economic and behavioral factors are associated to examination adherence, therefore, competing for a reduction in survival indicators, when disease is diagnosed in an advanced stage (SILVA et al, 2015).

In contrast, in order to avoid the low frequency of adequate samples, showing squamous and glandular cells, the collection must be representative of the squamocolumnar junction. The presence of endocervical and metaplasic cells in cytopathological smears has been considered as an indicator of collection quality, because it is the main source of pre-neoplasic lesions (UGHINI, 2016). Thus, the squamocolumnar junction is the part with the highest ocurrence of pre-neoplasic lesions, and endocervical and metaplasic cells must be present in samples for cytopathological examination; this works as an indicator for the lack of guidance and training of personell involved in the collection procedures (MONTEIO et al., 2009). Namely, it is possible to consider that the aim of screening for CC from examinations has being unsuccessful, besides the wrong use of public finantial resources and its impact in women’s health that failed to obtain information regarding their gynecological condition and possible treatments.

Furthermore, in case of an unsatisfactory result, the patient must redo the examination after a period of six to twelve months, since this timeframe may enable early CC diagnosis, and consequently the improvement of therapeutical response, in spite of the burden of the examination recurrence (BRASIL, 2013). This happens as one more disadvantage for the patients, considering that they will have to return to the health unit for a new sample collection for the smears, with possible expenses with displacement, work absence and discomfort at the collection moment. However, this return to the health unit does not happen quite often, and, therefore, these women lose the opportunity of an early diagnosis of possible harms, such as CC. The age range of 25 to 64 years, that predominated in the present study was also found in Aracaju, Sergipe (FERREIRA et al, 2015). These findings respond to the Brazilian MoH, and in this age range, precursor CC lesions are frequent (BRASIL, 2016). However, it is important to establish a minimal age for this age range.

According to the Brazilian MoH, women with no sexual activity record should not be screened for CC. However, it also points that screening may prevent the majority of cervical neoploasias, through the abnormal alterations identified in cervical cells (pre-neoplasias) (BRASIL, 2016), although these affirmations are contrary to the indication of screening onset at 25 years old. Thus, we understand that these arguments are controversial, favouring the suggestion of anticipating the age of onset of screening examinations.

According to Sasieni et al (2009), the incidence of CC in young women is low, and there are evidence of screening in women younger than 25 yeras old being less efficient than in older women. Similarly, in women younger than 25 years old LSIL are predominant, which regress spontaneously in most cases, and that annual examination does not increase significantly the protective effect of screening. Moreover, the Brazilian MoH emphasise that examinations out of target age range and recommended peridiocity could overload health services, and compromise access for those that have a precise indication. Namely, indiscriminate screening risk for women up to 24 years old would outweight its possible advantages (BRASIL, 2014; 2016).

Vetrano et al observed that international data showed a percentage of 12 to 45% of sexually active adolescents that had been screened for CC (VETRANO et al, 2007). Hence, the majority of these women sould have had indication for systematic CC screening, but not because they were young. In this regard, it is possible that many of these women may have precursor lesions, or other women needing treatment, but are not included in the current policies.

In addition, women that are not screened in the last two appointments show a four time higher risk of CC compared to those screened at the adequate period. Those that were not screened in na early appointment, but lately, also showed na elevated risk. Besides, being screened only with regular results has been associated to a CC from squamous cells risk reduction of 89%, compared to those that were not screened, but only 60% of reduction to adenocarcinoma (WANG et al, 2020).

Therefore, with the early onset of sexual activities, pregnancy and sexually transmited infections, we understand that this is a highly vulnerable group regarding the future development of CC. A study performed in Sao Paulo showed that on average, the onset of sexual activities happens at 14 years old, in contrast to the Brazilian MoH recommendations (CIRINO et al, 2010). Thus, there is a need for a policy revision regarding women’s health, in order to reduce the minimum age recommended for the onset of systematic CC screening, including younger women, for instance those younger than 20 years old.

Accordingly with the present results, the majority of women interviewed in a study in Rio Branco, Acre, were also Brown (89.1%) and had more than two sexual partners lifelong (70%) (PRADO et al, 2012). However, considering the social and historical context of Salvador county, the high number of Black/Brown women may not represent the interference of race/color on a higher risk for CC lesions in this sample.

In contrast to the findings from the present study, Borges et al (2012) observed a higher adherence to the preventive examination in those women who practiced physical activity (82.2%). A higher prevalence of non smokers in the present sample (94.7%) was also described in Porto Alegre, Rio Grande do Sul (75%). This low frequency is satisfactory, considering that smoking is a risk factor for CC development (UGHINI, 2016).

We observed a low proportion of vulvar and vaginal lesions in the present sample. Recurrent vulvovaginal infections are very frequent and has social and psicological consequences (NAMARTA et al, 2020). According to Curty et al (2020), in Fortaleza, Ceará, the reasons to cytopathological examinations are related to vulvovaginitis, which represents 70% of complaints in gynecological consultations (SOOST et al, 1991). However, in the present study, patients looked for examination preventively; that evidences the success of health education iniciatives focused on the present health unit recipients, in spite of inquiries regarding the quality of CC screening, considering weaknesses on sample collections that may contribute to a reliable diagnosis.

Moreover, we also observed in this study a high proportion of women without precursor CC lesions (88.7%), similarly to results observed by Silva in 2014 (86.8%). Tipically, we observed a high number of women presenting regular results for the samples analysed. That may indicate that those women that seek for extra health assistance may have their gynecological and reproductive health highly ensured.

Among possible cellular alterations, HSIL are truly precursor CC lesions, given its potential effect on disease progression. Therefore, its detection is considered as main focus of secondary prevention of CC (BRASIL, 2013). The present findings show that only 1.3% of examinations showed HSIL. However, if all examinations performed in the present study (n=1350) could have had a diagnosis, this proportion would represent a hypotherical number of 18 women with HSIL instead of two. Therefore, 16 women may have developed CC, whether or not they sough for early screening.

Finally, we point for the possible interference of the present sample size on the findings. This raises the need for studies with more robust samples. However, regardless of the sample size, we recognize the waste of public finantial resources on uneffective examinations, low effectiveness on CC screening by primary health care, and the need for higher investments on personell qualification and the revision of current clinical protocols for diagnosis, in order to have a better CC combat. A successfull experience was showed by Jakpbcynski et al, 2018 with personell qualification. They obtained a better sample, representation of required epithelium and examination results.

## FINAL CONSIDERATIONS

The high proportion of unsatisfactory samples described in the present results raises the need for investments on systematic qualification of personell involved in cytopathological examinations. That is relevant considering the identification of women that reported having many partners, therefore, being in higher risk for general gynecological alterations, including cervical cancer. In contrast, it requires additional costs and discomfort on recurrence of examinations, and may impact on the adherence for the systematic disease screening.

The proportion of unsatisfactory samples in cytopathological examinations is an indicator. It is related to the quality of collection, and also shows the proportion of samples that are considered inadequate or unsufficient for diagnosis, therefore, requiring another examination. Moreover, it allows the evaluation and formulation of qualification of human resources, in order to enhance finantial resources and avoid loss in women’s adherence to the examination.

On the other hand, the high frequency of young and sexually active women, that have more than one partner, and with a relatively low use of condoms, is associated with a higher risk for gynecological diseases. Therefore, the current minimal age for cervical cancer screening may be excluding these women. We understand the need for the implementation of protocols, in order to include these women in the screened group, and to standardise the collection and laboratory techniques, and to enhance and to streamline the diagnosis of precursor lesions for this type of cancer. Thus, this is possible if the State acomplish its duty of ensuring women the realization of their fundamental right for health, focusing on prevention and early detection of diseases and possible harms.

Thus, more studies regarding this topic are needed, especially those that explore the adequacy of collection and analysis of cytopathological examination, as an important tool on cervical cancer control, besides the effective prevention of its risk factors, in order to reduce the social and economic impact of this problem.

## Data Availability

The authors declare availability to present data mentioned in the manuscript.

## Notes

### Competing Interest Statement

The authors have declared no competing interest.

### Funding Statement

The authors declare that there was no financing for the execution of this work, nor did they receive payment or services from third parties for any aspect of the submitted work.

### Author Declarations

This study was approved by the Research Ethics Committee of the State University of Feira de Santana (No. 2,548,705).

## REFERENCES

Amaral R. G.; Manrique, E. J.C.; GuimarãEs, J. V.; Souza, P. J.; Mignoli, J. R.Q.; Xavier, A. P.; Oliveira, A. Influência da adequabilidade da amostra sobre a detecção das lesões precursoras do câncer cervical. Rev Bras Gincecol Obstet. 2008; 30(11):556–60.

Borges, M.F.de S.O, et al. Prevalência do exame preventivo de câncer do colo do útero em Rio Branco, Acre, Brasil, e fatores associados à não-realização do exame. Caderno de Saúde Pública, v. 28, n. 6, jun, Rio Branco, 2012.

BRASIL. Constituição (1988). Constituição da República Federativa do Brasil. Brasília, DF: Senado Federal: Centro Gráfico, 1988.

. Lei nº 8.080, de 19 de setembro de 1990. Lei Orgânica da Saúde. Dispõe sobre as condições para a promoção, proteção e recuperação da saúde, a organização e o funcionamento dos serviços correspondentes e dá outras providências. Brasília, set. 1990.

. Ministério da Saúde. Política Nacional de Atenção Integral à Saúde da Mulher: princípios e diretrizes. 1 ed., 2 reimpr. Brasília: Editora do Ministério da Saúde, 2011.

. Instituto Nacional de Câncer José Alencar Gomes da Silva. Monitoramento das ações de controle dos cânceres do colo do útero e de mama. Informativo Detecção Precoce, v. 3, n. 3, p. 1–12, ago/dez, 2012.

. Ministério da Saúde. Secretaria de Atenção à Saúde. Departamento de Atenção Básica. Controle dos cânceres do colo do útero e da mama / Ministério da Saúde, Secretaria de Atenção à Saúde, Departamento de Atenção Básica. – 2. ed. – Brasília: Editora do Ministério da Saúde, 2013.

. Instituto Nacional de Câncer José Alencar Gomes da Silva. Monitoramento das ações de controle dos cânceres do colo do útero e de mama. Informativo Detecção Precoce, v. 5, n. 1, p. 1–8, jan/abr, 2014.

. Instituto Nacional de Câncer José Alencar Gomes da Silva. Coordenação de Prevenção e Vigilância. Divisão de Detecção Precoce e Apoio à Organização de Rede. Diretrizes brasileiras para o rastreamento do câncer do colo do útero. 2. ed. rev. atual. – Rio de Janeiro: INCA, 2016.

. Ministério da Saúde. ABC do câncer: abordagens básicas para o controle do câncer / Instituto Nacional de Câncer José Alencar Gomes da Silva; organização Mario Jorge Sobreira da Silva. – 3. ed. rev. atual. – Rio de Janeiro: Inca, 2017.

Cirino, F.M.S.B. et al. Conhecimento, atitude e práticas na prevenção do câncer de colo uterino e hpv em adolescentes. Escola Anna Nery, v. 14, n. 1, p. 126–134, Mar., Rio de Janeiro, 2010.

Curty, G. et al.. O papel do microbioma cervicovaginal na gênese e como biomarcador de neoplasia intraepitelial cervical pré-maligna e câncer cervical invasivo. Jornal internacional de ciências moleculares, v. 21, n. 1, p. 222. 2019.

Ferreira, J.E.L. et al.. Perfil da população atendida em um consultório de atendimento integral à saúde da mulher. Ciências Biológicas e da Saúde, v. 3, n. 1, p. 127–140, out., Aracajú, 2015.

GONTijo, R.C. et al.. Avaliação de métodos alternativos a citologia no rastreamento de lesões cervicais: detecção de DNA-HPV e inspeção visual. Rev. Bras. Ginecol. Obstet. v. 26, n. 4, maio, Campinas, 2004.

Jakobczynski J, Frighetto M, Perazzoli M, Dambrós BP, Dallazem B, Kirschnick A. Capacitação dos profissionais de saúde e seu impacto no rastreamento de lesões precursoras do câncer de colo uterino. RBAC. 2018;50(1):80–5.

Lapin, G. A. Derchain, S. F. M; Tambascia, J. Comparação entre a colpocitologia oncológica de encaminhamento e a gravidade das lesões cervicais intra-epiteliais. Revista de Saúde Publica, v. 34, n. 2, abril, Campinas, 2000.

LeitãO, N.M.A. et al.. Avaliação dos laudos citopatológicos de mulheres atendidas em um serviço de enfermagem ginecológica. Revista Mineira de Enfermagem, v.12, n. 4, out./dez., Ceará, 2008.

Namarta, K.; Jatinder S.; Manpreet, Kl. Microbiota in vaginal health and pathogenesis of recurrent vulvovaginal infections: a critical review. Ann Clin Microbiol Antimicrob. v. 19, n. 1, jan., 2020.

Prado, P.R. et al.. Caracterização do Perfil das Mulheres com Resultado Citológico ASCUS/AGC, LSIL e HSIL segundo Fatores Sociodemográficos, Epidemiológicos e Reprodutivos em Rio Branco - AC, Brasil. Revista Brasileira de Cancerologia, v. 58, n. 3, p. 471–9, Acre, 2012.

Santos, K.M. Distribuição espacial da mortalidade por câncer de colo de útero no Brasil, 1996 a 2009: eco-cuidado de enfermagem. 2011, 110 f. Dissertação (Mestrado em Enfermagem) - Universidade Federal do Estado do Rio de Janeiro. Rio de Janeiro, 2011.

Sasieni, P. et al.. Effectiveness of cervical screening with age: population based case-control study of prospectively recorded data. British Medical Journal, v. 339, n. 2968, p. 328, 2009.

Silva, P.V.; AraúJo, A.; AraúJo, M.R.N. Análise da cobertura do exame citopatológico do colo do útero no município de Doresópolis-MG. Revista de Enfermagem do Centro Oeste Mineiro, v. 1, n. 2, p. 154–163, 2011.

Silva, M. A. S. et al.. Fatores relacionados a não adesão à realização do exame de Papanicolau. Revista Rene, v. 16, n. 4, p. 532–539, 2015.

Soost, H.J. et al.. The validation of cervical cytology: sensitivity, specificity and predictive values. Acta Cytol, v. 35, p. 8–13, 1991.

Ughini, S.F.O. Importância da qualidade da coleta do exame preventivo para o diagnóstico das neoplasias glandulares endocervicais e endometriais. Departamento de Análises, Universidade Federal do Rio Grande do Sul. Porto Alegre 2016.

Vetrano G, Lombardi G, Di Leone G, Parisi A, Scardamaglia P, Pate G, et al. Neoplasia intraepitelial cervical: fatores de risco para persistência e recorrência em adolescentes. Eur J Gynaecol Oncol. 2007; 28 (3): 189–92.

Wang, J., ElfströM, K. M., Andrae, B., Nordqvist Kleppe, S., Ploner, A., Lei, J., Dillner, J., SundströM, K., SparéN, P. Cervical cancer case-control audit: results of the routine evaluation of a national cervical screening program. International Journal of Cancer, v. 146, n. 5, p. 1230–1240, mar., 2020. https://doi.org/10.1002/ijc.32416

